# Evidence for increased breakthrough rates of SARS-CoV-2 variants of concern in BNT162b2 mRNA vaccinated individuals

**DOI:** 10.1101/2021.04.06.21254882

**Authors:** Talia Kustin, Noam Harel, Uriah Finkel, Shay Perchik, Sheri Harari, Maayan Tahor, Itamar Caspi, Rachel Levy, Michael Leschinsky, Shifra Ken Dror, Galit Bergerzon, Hala Gadban, Faten Gadban, Eti Eliassian, Orit Shimron, Loulou Saleh, Haim Ben-Zvi, Doron Amichay, Anat Ben-Dor, Dana Sagas, Merav Strauss, Yonat Shemer Avni, Amit Huppert, Eldad Kepten, Ran D. Balicer, Doron Nezer, Shay Ben-Shachar, Adi Stern

## Abstract

The SARS-CoV-2 pandemic has been raging for over a year, creating global detrimental impact. The BNT162b2 mRNA vaccine has demonstrated high protection levels, yet apprehension exists that several variants of concerns (VOCs) can surmount the immune defenses generated by the vaccines. Neutralization assays have revealed some reduction in neutralization of VOCs B.1.1.7 and B.1.351, but the relevance of these assays in real life remains unclear. We performed a case-control study that examined the distribution of SARS-CoV-2 variants observed in infections of vaccinated individuals (“breakthrough cases”) and matched infections of unvaccinated individuals. We hypothesized that if there is lower vaccine effectiveness against one of the VOCs, its proportion among the breakthrough cases should be higher than among unvaccinated cases. Our results show that vaccinees that tested positive at least a week after the second dose were indeed disproportionally infected with B.1.351, as compared with unvaccinated individuals (odds ratio of 8:1). Those who tested positive between two weeks after the first dose and one week after the second dose, were disproportionally infected by B.1.1.7 (odds ratio of 26:10), suggesting reduced vaccine effectiveness against both VOCs at particular time windows following vaccination. Nevertheless, the B.1.351 incidence in Israel to-date remains low and vaccine effectiveness remains high among those fully vaccinated. These results overall suggest that vaccine breakthrough infection may be more frequent with both VOCs, yet a combination of mass-vaccination with two doses coupled with non-pharmaceutical interventions control and contain their spread.

---

Mass vaccination against severe acute respiratory syndrome coronavirus 2 (SARS-CoV-2) is currently underway worldwide, providing hope that the COVID-19 pandemic may soon be mitigated. In Israel, vaccination commenced on December 20, 2021, primarily with the BNT162b2 mRNA vaccine, and by mid-March 2021, over 80% of the eligible population (all individuals 16 years old and above) were vaccinated with at least one dose. In clinical trials, the BNT162b2 mRNA vaccine was shown to be 95% efficacious in preventing symptomatic disease; a similarly high protective effectiveness has also been found in real-world settings in Israel [1, 2]. However, concerns have emerged regarding the effectiveness of vaccines against various SARS-CoV-2 strains. In particular, three strains have been recently defined as “variants of concern” (VOC) by the world health organization (WHO; www.who.int): the B.1.1.7 strain (originally detected in the United Kingdom), the B.1.351 strain (originally detected in South Africa), and the P.1 strain (originally detected in Brazil). Accumulating evidence suggests that the B.1.1.7 strain spreads more rapidly than the original circulating strain and is accompanied by increased mortality rates [3, 4].

Concerns have emerged that the B.1.351 and P.1 strains are able to overcome previous immunity to SARS-CoV2 [5, 6], yet the evidence has been somewhat mixed. Using engineered viruses and/or sequences, laboratory studies have shown that neutralization of B.1.1.7 by BNT162b2 vaccine-elicited sera was either similar or slightly reduced as compared to neutralization of the original circulating strain. Conversely, a significant reduction in neutralization of B.1.351 was observed [7-11], while other studies suggested neutralization remained relatively high against both B.1.1.7 and B.1.351 [12]. T-cell responses, which are not captured by neutralization studies, were also shown to remain stable against these variants following vaccination [13]. Thus, it remains unknown whether VOCs can perform BNT162b2 vaccine breakthrough in real world settings, in which the vaccine maintain both antibody and T-cell responses.

Here we tested the hypothesis that the B.1.1.7 and B.1.351 strains are able to overcome BNT162b2 mRNA vaccine protection. To this end, we identified individuals with documented SARS-CoV-2 infection – symptomatic or asymptomatic (hereby denoted as carriers) amongst members of the Clalit Health Services (CHS), the largest health care organization in Israel, which insures 4.7 million patients (53% of the population). We focused on vaccinated carriers, divided into two categories: (a) individuals who had a positive PCR test that was performed between 14 days after the 1^st^ dose and a week after the 2^nd^ dose (denoted as partial effectiveness, PE), and (b) individuals who had a positive PCR test that was performed at least one week after the second vaccine dose (denoted as full effectiveness, FE). The latter was chosen to match our previous study that showed very high real-world vaccine effectiveness in Israel, using this particular criterion [1]. Each vaccinee (case) was matched with an unvaccinated carrier (control) with similar demographic characteristics (date of PCR, age, sex, ethnic sector, and geographic location) to reduce bias associated with differential exposure (Methods). Next, we collected RNA from the PCR samples and performed complete viral genome sequencing for 813 samples from different individuals, consisting of 149 pairs of FE-controls, 247 pairs of PE-controls and additional samples whose match did not undergo successful sequencing (see below) (Table 1, Table S1, Fig. S1). When examining the results, it became evident that B.1.1.7 was the predominant strain of virus in Israel over the entire sampling period, increasing in frequency over time (Fig. 1A). Conversely, the B.1.351 strain was at an overall frequency of less than 1% in our sample, confirming previous reports (Fig. 1B) [14]. No other variants of concern or variants of interests, as defined by the WHO, were found in our sample (Fig. S2). We collectively denote all other lineages found as wild-type (WT) (Methods). Moreover, we did not find evidence for the increased presence of any additional mutations that are not lineage defining mutations of B.1.1.7 or B.1.351.

**Table 1.** Demographic statistics on paired cases and controls sequenced herein. Absolute counts are shown, relative proportions are in brackets.

|  |  | Control FE vaccinee | Control PE vaccinee | FE vaccinee | PE vaccinee |
| --- | --- | --- | --- | --- | --- |
|  | Number | 149 | 247 | 149 | 247 |
| Age group | 0-19 | 2 (1.3) | 4 (1.6) |  | 1 (0.4) |
|  | 20-29 | 20 (13.4) | 31 (12.6) | 5 (3.4) | 31 (12.6) |
|  | 30-39 | 32 (21.5) | 59 (23.9) | 12 (8.1) | 48 (19.4) |
|  | 40-49 | 33 (22.1) | 59 (23.9) | 31 (20.8) | 64 (25.9) |
|  | 50-59 | 22 (14.8) | 55 (22.3) | 24 (16.1) | 53 (21.5) |
|  | 60-69 | 24 (16.1) | 25 (10.1) | 30 (20.1) | 32 (13.0) |
|  | 70-79 | 9 (6.0) | 10 (4.0) | 24 (16.1) | 13 (5.3) |
|  | 80-89 | 7 (4.7) | 4 (1.6) | 22 (14.8) | 5 (2.0) |
|  | 90+ |  |  | 1 (0.7) |  |
| Sex | F | 87 (58.4) | 152 (61.5) | 81 (54.4) | 152 (61.5) |
|  | M | 62 (41.6) | 95 (38.5) | 68 (45.6) | 95 (38.5) |
| District | Dan | 25 (16.8) | 22 (8.9) | 23 (15.4) | 22 (8.9) |
|  | South | 3 (2.0) | 5 (2.0) | 3 (2.0) | 5 (2.0) |
|  | Haifa | 45 (30.2) | 70 (28.3) | 45 (30.2) | 70 (28.3) |
|  | Jerusalem | 29 (19.5) | 71 (28.7) | 29 (19.5) | 71 (28.7) |
|  | Center | 23 (15.4) | 28 (11.3) | 23 (15.4) | 28 (11.3) |
|  | North | 7 (4.7) | 22 (8.9) | 7 (4.7) | 22 (8.9) |
|  | Sharon-Shomron | 7 (4.7) | 19 (7.7) | 9 (6.0) | 19 (7.7) |
|  | Tel-Aviv | 10 (6.7) | 10 (4.0) | 10 (6.7) | 10 (4.0) |
| Sector | General Jewish | 117 (78.5) | 163 (66.0) | 117 (78.5) | 163 (66.0) |
|  | Jewish Orthodox | 11 (7.4) | 28 (11.3) | 11 (7.4) | 28 (11.3) |
|  | Non-Jewish | 21 (14.1) | 56 (22.7) | 21 (14.1) | 56 (22.7) |
| Variant | B.1.1.7 | 138 (92.6) | 206 (83.4) | 134 (89.9) | 221 (89.5) |
|  | B.1.351 | 1 (0.7) | 1 (0.4) | 8 (5.4) | 1 (0.4) |
|  | WT | 10 (6.7) | 40 (16.2) | 7 (4.7) | 25 (10.1) |
| Vaccine status | Non-vaccinated | 149 (100.0) | 247 (100.0) |  |  |
|  | 14-20 days from 1st dose |  |  |  | 133 (53.8) |
|  | 21-28 days from 1st dose |  |  |  | 95 (38.5) |
|  | 28+ days from 1st dose |  |  |  | 19 (7.7) |
|  | 7-13 days from 2nd dose |  |  | 73 (49.0) |  |
|  | 14-20 days from 2nd dose |  |  | 30 (20.1) |  |
|  | 21+ days from 2nd dose |  |  | 46 (30.9) |  |

**Figure 1.**
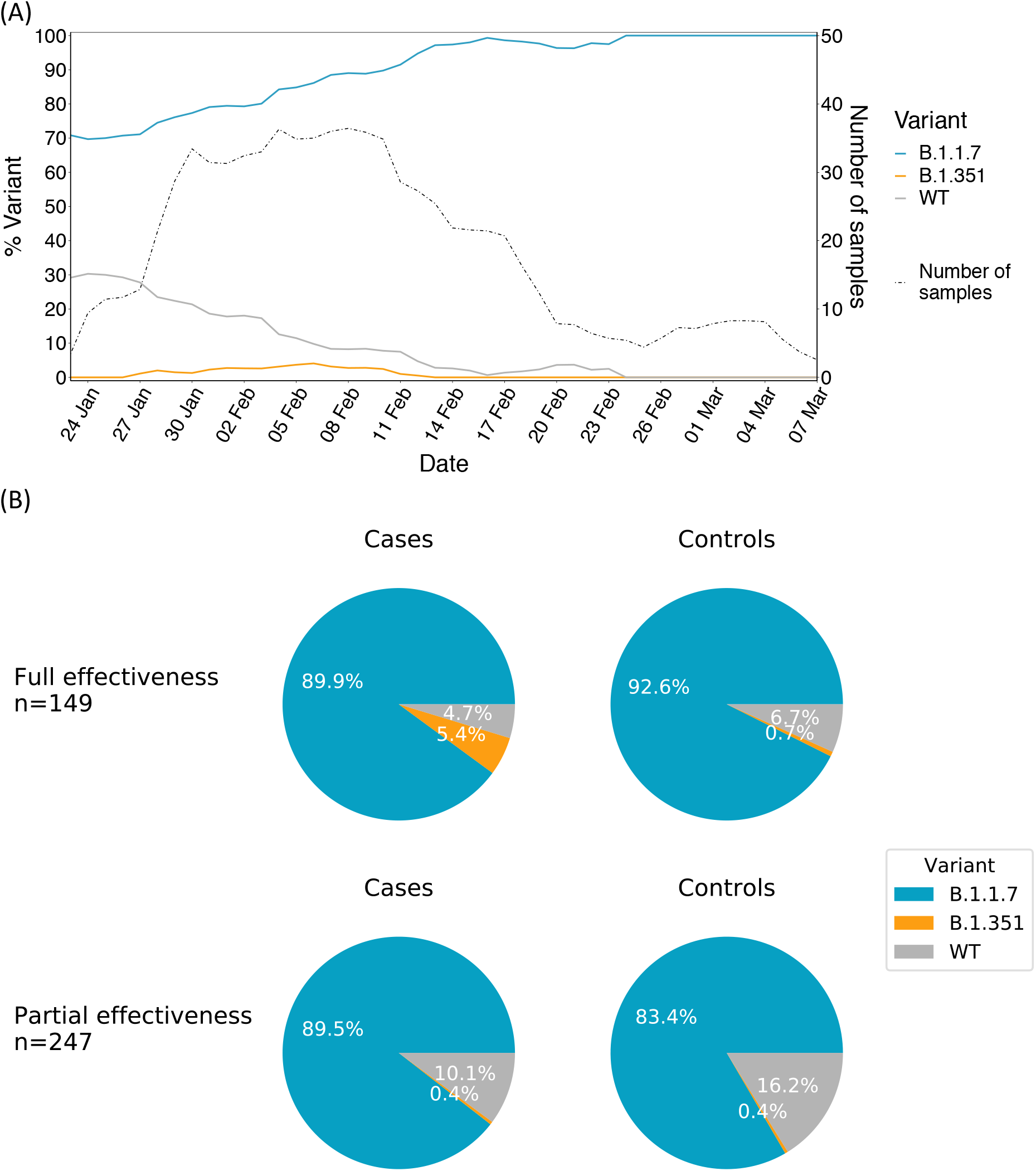
Variant frequencies of SARS-CoV-2 positive samples. (A) Variant frequencies are shown across the time of the study, including the number of samples collected throughout the study. All values were calculated by averaging over a sliding window of seven days. (B) Breakdown of variant frequencies based on the four groups of this study: pie charts display the proportion of each variant (B.1.1.7, B.1.351, WT) for paired vaccinated cases versus non-vaccinated controls separated by effectiveness (full effectiveness and partial effectiveness, as defined in the main text), with cases on the left and their associated control on the right.

We next analysed our paired set of vaccinated and non-vaccinated carriers, using a stringent method of lineage assignment for each viral sequence (Methods). Based on previous results from neutralization assays, we hypothesized that B.1.1.7 may be slightly vaccine-resistant as compared to WT, whereas B.1.351 may be more vaccine-resistant when compared to both B.1.1.7 and WT. Under this hypothesis of ordered resistance, we performed our statistical analyses first on the B.1.1.7 strain, while excluding B.1.351 strains (to avoid obscuring a potential signal), and then compared the B.1.351 with the B.1.1.7 and WT variants combined (Fig. 2). Our null model was that under a hypothesis of equal effectiveness of the vaccine against all variants, we would expect each variant to be represented at a similar proportion in the vaccinated groups (FE & PE), as compared to their matched control groups.

**Figure 2.**
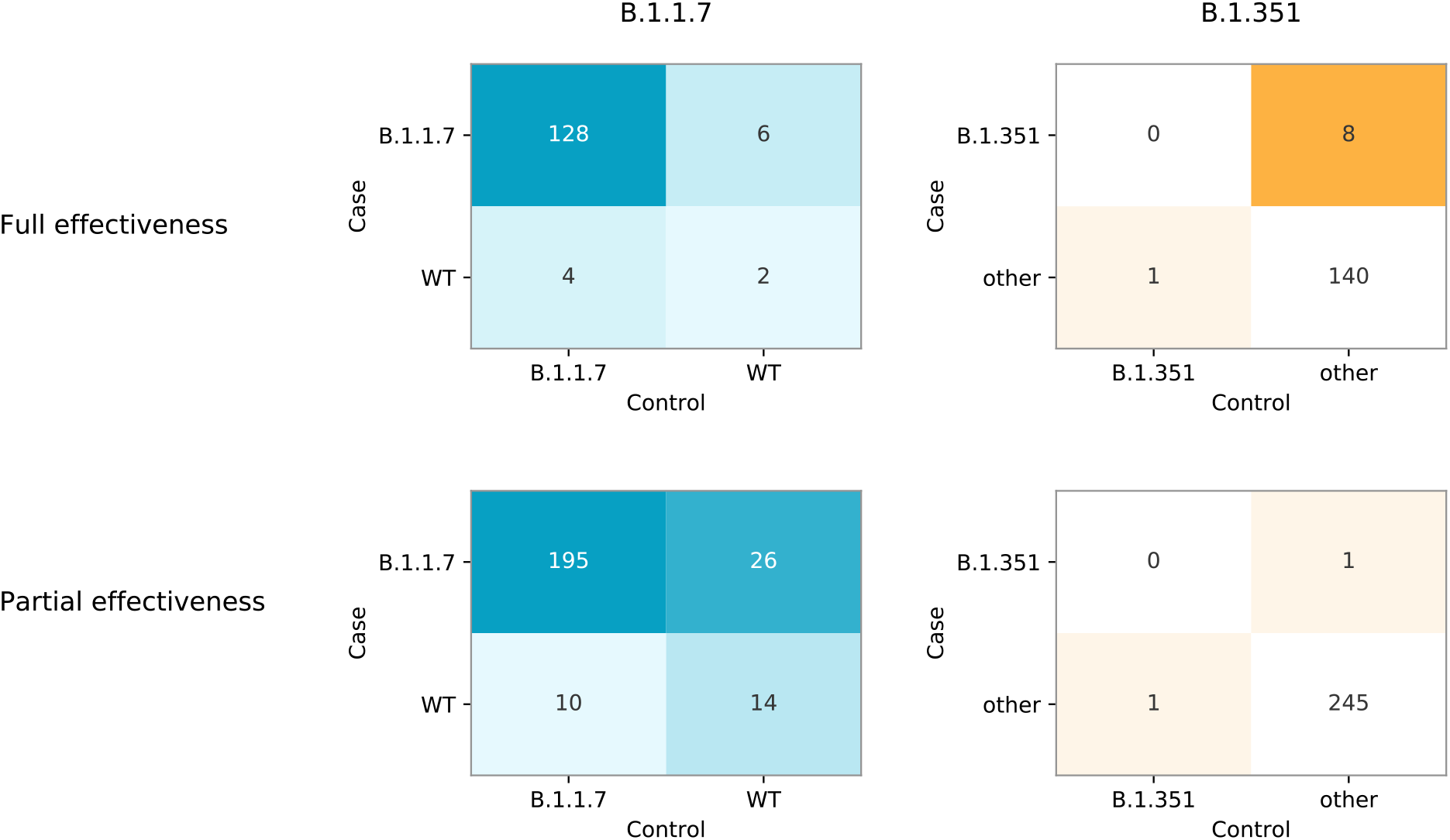
Results of matched vaccinated cases and non-vaccinated controls separated by effectiveness and variant of concern. In each table, a cell reflects the number of pairs concordant (upper left and lower right) or discordant (upper right or lower left) for a given variant. The left panel focuses on the comparison between B.1.1.7 and WT (pairs with B.1.351 were removed), whereas the right panel focuses on comparing B.1.351 and either WT or B.1.1.7 (denoted collectively as “other”). Of note, the McNemar test focuses on a comparison of discordant samples only.

No statistically significant difference was observed in the rates of B.1.1.7 infection in FE cases versus unvaccinated controls (odds ratio [OR] of 6:4, one-sided exact McNemar test, p=0.38), but a significantly higher proportion of B.1.351 was observed in FE cases vs. unvaccinated controls (OR of 8:1, one-sided exact McNemar test, p=0.02). Of note, about half of FE cases tested positive on days 7-13 days after the second dose, and about half tested positive 14 days or more after the second dose (Table 1). However, all eight FE cases with B.1.351 were infected 7-13 days after the second dose.

On the other hand, a significantly higher rate of B.1.1.7 was observed in PE cases vs. unvaccinated controls (OR of 26:10, one-sided exact McNemar test, p=0.006). For B.1.351 in the PE category, the sparsity of data (one infection in each category) precluded statistical analysis (Fig. 2). A conditional logistic regression was further performed on the PE B.1.1.7 data (since more data was available in this category), supporting the previous analysis: an OR of 2.4 was observed (95% confidence interval of 1.2 to 5.1). Age was included in the regression and was found to be a non-significant confounder, suggesting that its possible role in propensity for infection by a specific strain was corrected through our matching scheme.

To test if our sampling scheme was biased, we reconstructed a phylogenetic tree of all the sequenced samples together with additional available sequences from Israel, and observed that vaccinated and unvaccinated samples were highly interspersed along the tree (Fig. 3) ruling out strong biases in sampling.

**Figure 3.**
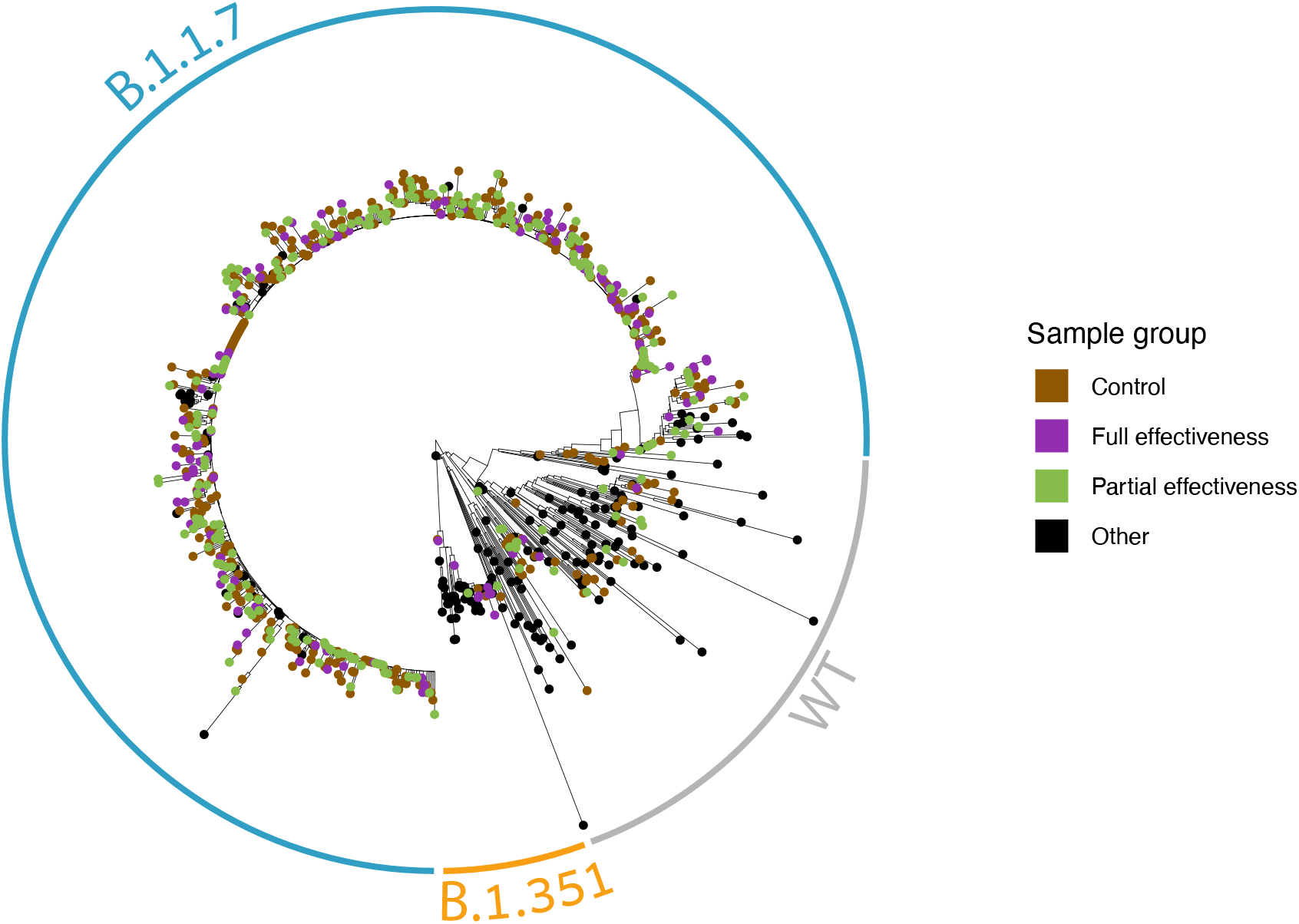
A maximum-likelihood phylogenetic tree of Israeli SARS-CoV-2 samples including those sequenced herein. Vaccinees are coloured in violet or green, non-vaccinees are coloured in brown, and black sequences are publicly available sequences from Israel (marked as “other”). Clades composed of the B.1.1.7, B.1.351 and WT sequences are encircled in blue, orange and grey, respectively.

Finally, we noted an additional two B.1.351 sequences, consisting of one FE case and one PE control, where the sequencing of the matched pair did not undergo successful sequencing, most often due to high Ct (low viral load). Importantly, these pairs would either leave our conclusions unchanged, or would increase the odds-ratio in favour of the B.1.351 in the FE category (Fig. S3), strengthening the results reported above. With regards to B.1.1.7, we found a total of 28 non-paired sequences, once again since a control or case yielded unreliable sequencing. These sequences might change the significance of our results with regards to B.1.1.7 but would not change the trend we found for this variant (Fig. S3).

## Discussion

Our results show that there is an increased incidence of VOC B.1.351 in vaccine breakthrough infections in fully vaccinated individuals with BNT162b2, and increased incidence of VOC B.1.1.7 in partially vaccinated individuals (Figs. 2,3). These results are generally aligned with those from in vitro neutralization assays that have shown a large reduction in neutralization against B.1.351, and little to no reduction against B.1.1.7 in fully vaccinated individuals [7-11]. Overall, our data also suggests that serum-based neutralization studies that take into account individual monoclonal antibody responses may provide a good proxy for real life protection in the case of SARS-CoV-2 [15]. Although this remains to be tested in a more widespread manner, it suggests that neutralization studies may be valid as a prompt first step prior to the establishment of real-world studies in the case of the emergence of new SARS-CoV-2 VOCs.

The power of our approach stems from the combination of real-world evaluation with the stringent case-control matching strategy employed, allowing us to rule out that a high proportion of a given variant was due to a confounding effect. However, it is still possible that other confounding effects were present and were not controlled for, i.e. behavioural effects among vaccinees. Additionally, sequencing limitations prevented us from sequencing very low viral load samples (Methods), and thus the focus of our study was on vaccinees who generated higher viral loads. However, it has been shown that cases with low viral load may be a lesser concern from a public health perspective, as they are associated with less symptoms and decreased transmission [16]. Finally, our FE cohort is based on infections documented seven or more days post the second vaccine dose (Table 1). Some subjects in this cohort may have been infected before the immunity from the boost was fully established, and it is thus possible that enhanced immunity from the boost, which develops over time [17], may more effectively prevent infection with the B.1.351 variant. Notably, when focusing on the eight B.1.351 cases in the FE group, all tested positive during days 7-13 post the second dose, and none tested positive in days 14+ post the second dose. This observation suggests that increased breakthrough of B.1.351 in our cohort occurs mainly in a limited time window post vaccination. Further research is required to clarify these key issues.

The main caveat of our study was the small sample size of both the WT and B.1.351 variants. These small samples sizes are a product of (a) the dramatic increase in frequency of the B.1.1.7 variant (first detected in Israel in mid-December 2020) (Fig. 1A), and (b) the low frequency of the B.1.351 variant in Israel [14]. In fact, in our latest samples obtained in late February and early March 2021, we noted fixation of the B.1.1.7 variant, but this interpretation requires caution as our sample size was low (Fig. 1A). Furthermore, caution is required from over-interpreting the odds ratios obtained, as the absolute numbers we found, in particular for B.1.351 infections, are very small.

Our study design was not intended to deduce vaccine effectiveness against either variant, since we observe VOCs conditioned on infection, and do not measure absolute infection rates in the vaccinated or control population. Thus, we can only cautiously speculate on vaccine effectiveness against the B.1.1.7 and B.1.351 strains from these dates. Previous real-world work has shown a very high effectiveness of the BNT162b2 vaccine starting a week after the second dose [1]. During the period of the latter study, B.1.1.7 likely rose to a high frequency, suggesting high vaccine effectiveness also against this strain. However, our current study may suggest a somewhat lower protection B.1.1.7 in the first weeks after the first vaccine dose. As some countries opt to increase the gap from first to second BNT162b2 vaccine from the recommended 3 weeks to longer period [18], it is important to carefully assess whether this delay impacts vaccine effectiveness against the B.1.1.7 strain.

From a biological point of view, the breakthrough cases observed in this study might either be due to immune evasion of both strains, or the ability of B.1.17 to create higher viral loads [3]. Given the low frequency of B.1.351 across time (Fig. 1A) [14], our results overall suggest that selection does not strongly favour the B.1.351 variant in the particular conditions in Israel. In view of this low frequency of B.1.351 (across all groups of study herein, including the FE category; Fig. 1B), we suggest that there may be higher rates of vaccine breakthrough with B.1.351, but it is possible that (a) vaccine effectiveness coupled with enacted non-pharmaceutical interventions remain sufficient to prevent its spread, and/or (b) B.1.1.7 outcompetes B.1.351, possibly due to its high transmission rate [3]. Our results emphasize the importance of tracking viral variants in a rigorous framework and of increasing vaccination, which we conclude is the safest and most effective means of preventing the onwards spread of B.1.351 and other possible future VOCs.

## Methods

### Ethics statement

The study was approved by the CHS institutional review board (IRB #0016-21-COM2) and was exempt from the requirement for informed consent. The study was further approved by the Tel-Aviv University ethics committee (0002706-1).

### Sample matching

Data for this study were obtained from CHS’s data repositories (Fig. S1). The study population consisted of individuals who tested positive for SARS-CoV-2 by RT-PCR test from six major CHS testing labs located throughout Israel. Subjects with positive PCR were then classified into one of two groups: (a) controls who were not vaccinated prior to the positive PCR result, (b) cases that were vaccinated at least two weeks prior to the PCR result. Cases were further divided into two additional sub-groups: (b1) subjects who obtained the first dose of vaccination, at least 14 days but earlier than a week after the 2^nd^ dose prior to the PCR result, denoted as the partial effectiveness PE sub-group, and (b2) subjects who obtained the 2^nd^ vaccine’s dose one week or more prior to the PCR results, denoted as the full effectiveness FE sub-group. Next, each case was matched to a control using six parameters: date of sampling for PCR (+/-three days), sex, age (+/-10 years), municipality of residence, geographical district of residence, and sector. In preliminary analyses, we noted that matching often failed for FE samples due to their small sample size, as well as due to increasing proportions of vaccinated individuals in older age categories across time, in line with the vaccine rollout policy in Israel. To increase FE matching, we enforced matching on date of PCR sampling, but allowed for four out of five matches in the remaining parameters. We found that the failed parameter match was most often age, sex, or municipality. We note that ten control samples served as controls for both an FE and a PE sample. Table 1 summarizes statistics on the parameters used for matching and other parameters for the various groups of our sample.

### Obtaining RNA samples and sequencing

Following matching, RNA from cases and controls was obtained from the main testing laboratories of CHS, with one major limitation: only samples with Ct values of 33 or lower were collected. Dates of samples ranged from January 23, 2020 and March 07 2021 (Fig. S4). Full genome sequencing of SARS-CoV-2 was performed based on the Artic protocol with a V3 primer set (https://artic.network/ncov-2019), with slight modifications detailed below. Briefly, reverse transcription and multiplex PCR was performed in two amplicon pools, and NEBNext Multiplex Oligos for Illumina were ligated to allow for sequencing. All samples were run on an Illumina Miseq using 250-cycle V2 kits at either the Technion Genome Center (Israel) or at the Genomics Research Unit at Tel Aviv University (Israel). We and others have previously noted amplicon dropout of amplicons 74 and 76 [19], both of which cover the Spike gene, and in particular some of the lineage-defining mutations of B.1.1.7 and B.1.351 (such as E484K and N501Y). To increase the sequencing yield of these amplicons we doubled the primer concentrations of both amplicons in our primer pool and lowered the annealing-extension temperature to 63°C.

### Bioinformatic analysis and lineage assignment

Sequencing reads were trimmed using pTrimmer, a multiplexing primer trimming tool [20], and then aligned to the reference genome of SARS-CoV-2 (GenBank ID MN908947) using our AccuNGS pipeline [21] that is based on BLAST [22], using a particular stringent e-value of 10^−30^. We then set out to determine the consensus sequence of each sample. Typically studies often report a majority rule consensus sequences, i.e, the consensus base at each position in the genome is the base that most reads (>50%) support. However, the biological meaning of variable positions where more than one base is observed is complex, especially if such positions are abundant: they may indicate within-host variation, they may indicate multiple genotype infection, they may indicate sample contamination, and they may indicate sequencing errors. To overcome these limitations, we constructed two consensus sequences for each sample, one based on majority-rule, and a more strict consensus sequence where we required >=80% of reads to support a given base. Bases with lower support were assigned an “N” (ambiguous base). In both types of consensus sequencing assignments, we required sequencing coverage of at least ten reads. Finally, we used the Pangolin software (github.com/cov-lineages/pangolin) to assign lineages for each consensus sequence using the Pango nomenclature [23], which requires that at least 50% of bases sequenced are unambiguous. After verifying the type of lineages we obtained, we labelled all consensus sequences as either “B.1.1.7”, “B.1.351”, or “WT”. Samples for which Pangolin labels of the strict and majority rule consensuses did not coincide were discarded. Thirty two pairs in which one sample did not undergo successful sequencing were discarded from the paired analyses (but see Fig. S3). The unpaired successful samples were however included in the variant frequencies across time analysis (Fig 1A).

### Data availability

All sequences were uploaded to GISAID. Submission of the raw sequencing data to Sequence Read Archive (SRA) is pending. A list of all sequence accession numbers used in this study will be available in Table S1 (pending).

### Statistical analysis

For all primary analyses a one-sided paired (exact) McNemar’s test was used in order to compare breakthrough of a variant in partially or fully vaccinated individuals. Notably, this test does not allow reporting a confidence interval for the inferred odds-ratio. For the analysis of B.1.351, all other variants were defined as the reference group, while for the B.1.1.7 analysis we excluded any paired observation that included B.1.3.5 (assuming ordinality of breakthrough), while any other variant was defined as the reference. Conditional logistic regressions were used as a sensitivity analysis in order to include age as a possible confounder in case that matching was not sufficient, under the assumption that it was sometimes only partially mediated through matching. All analyses were conducted with the use of R software version 4.03 and the survival and exact2×2 packages.

### Phylogenetic analysis

All Israeli sequences available on GISAID (https://www.gisaid.org/) from August onwards were downloaded, focusing on high quality sequences with 10% or less ambiguous sites. Of these sequences, due to computational intensity, we sampled the most distant 100 WT sequences and 50 B.1.1.7 sequences, and included all available B.1.351 sequences. The reference genome sequence (MN908947.3) was added on as well and these sequences were combined with sequences from this study that contained at most 10% ambiguous sites. Alignment was performed using Mafft [24] with default parameters. Next, a maximum-likelihood phylogeny was reconstructed using PhyML [25] with default parameters as well and the tree was rooted using the MN908947.3 sequence from Wuhan.

## Data Availability

All sequences were uploaded to GISAID.

## Contributions

T.K. and N.H performed the bioinformatics analyses

S.P. UF, E.K and M.L performed the clinical data analyses

A.H. provided consultation on study design and statistical analyses

U.F, S.P., T.K., N.H. and A.S performed the statistical analyses

T.K, N.H, U.F, S.P., E.K., S.BS and A.S. drafted the paper

T.K, N.H, S.H., M.T, and R.L coordinated clinical sample collection and sequencing

S.H. and M.T. performed library preparation for sequencing

S.K.D., G.B., H.G., F.G., E.E., O.S., L.S., H.B., D.A., A.B., D.S., M.S., and Y.S.A. contributed clinical samples

R.M., T.K, N.H, S.P, E.K, D.N., R.D.B, S.BS and A.S. conceived and coordinated the study

A.S., D.N, and S.BS. supervised and led the study

## Acknowledgements

We would like to thank Ron Milo, Naama Kopelman, Yonatan Woodbridge, David Burstein and Tzachi Hagai for their advice and discussions, Sydney Krispin for editing support, and Omer Tirosh and Moran Meir for extensive technical support. We would also like to thank the Technion Genome Center for rapid sequencing services. This study was supported by an ERC starting grant 852223 (RNAVirFitness), by an Israeli Science Foundation grant 3963/19, and by kind donations from the Millner and AppFlyer foundations. This study was supported in part by fellowships to TK, NH, and SH from the Edmond J. Safra Center for Bioinformatics at Tel-Aviv University.

