## Supplementary figures for "Evidence for increased breakthrough rates of SARS-CoV-2 variants of concern in BNT162b2 mRNA vaccinated individuals"

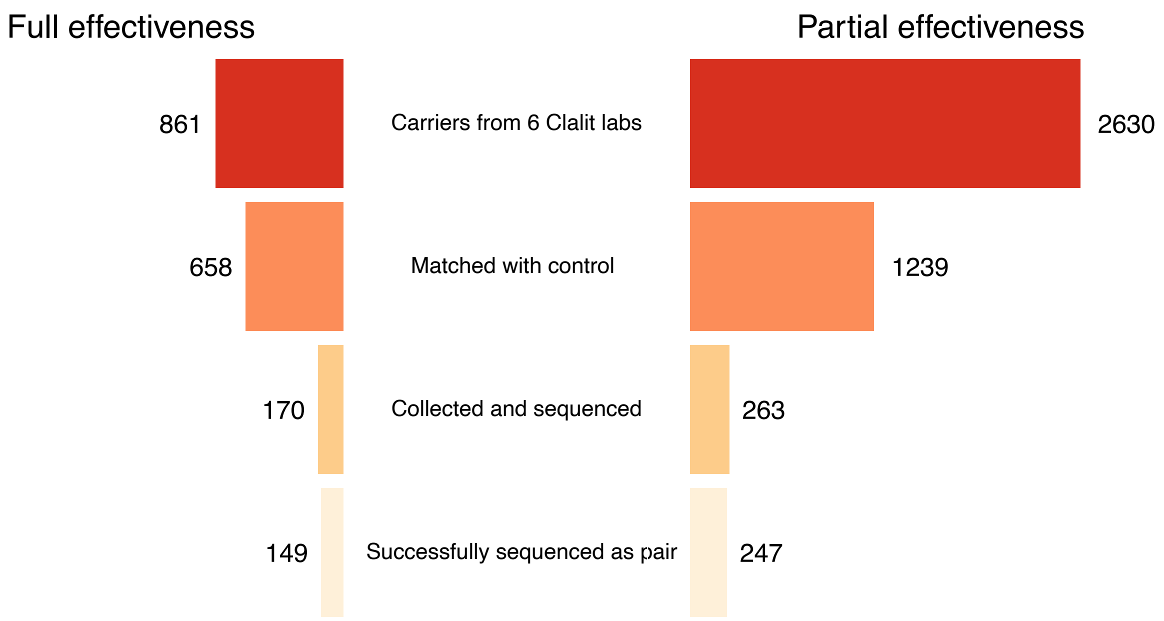


**Fig. S1. Number of vaccinated carriers, from initial identification to successful sequencing**. Matching with a control often failed due to two main reasons: (a) increasing proportions of the population were vaccinated, leaving smaller numbers of unvaccinated controls, (b) our strict requirement for matching of geographic location.

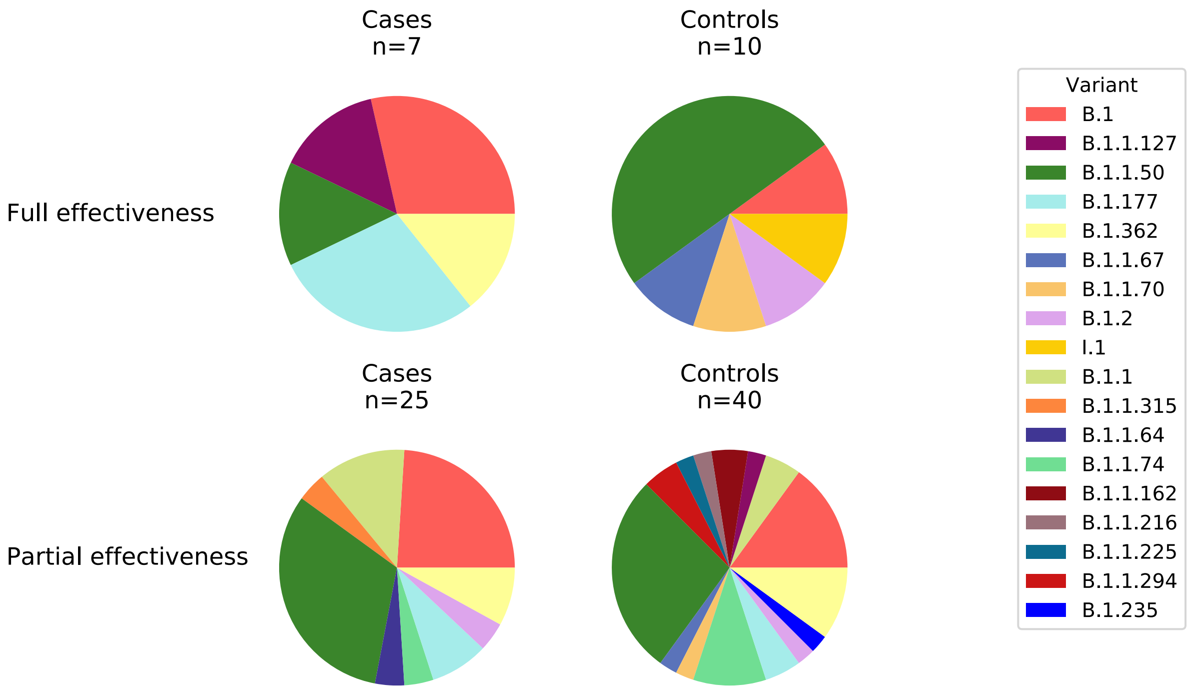


**Fig. S2. WT variants (lineages) found in this study, partitioned into the four groups studied herein.** Lineages were inferred based on strict consensus sequences (Methods).

(A)

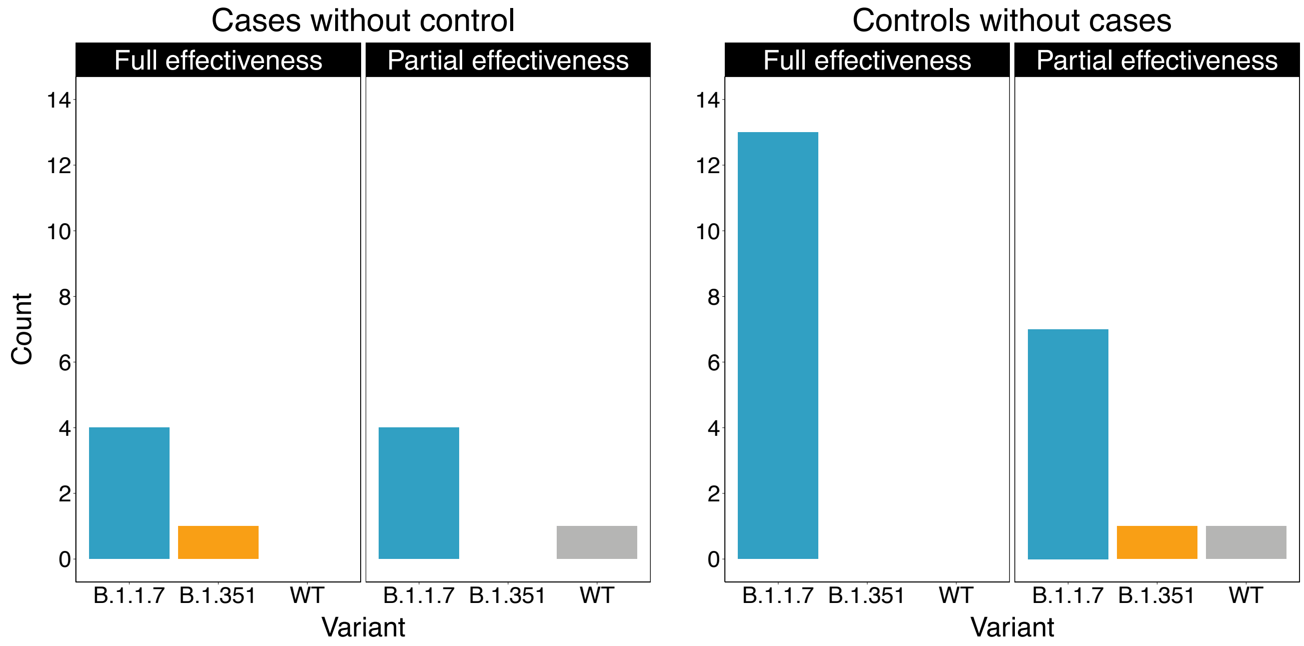


(B)

Scenario that most strengthens our results:

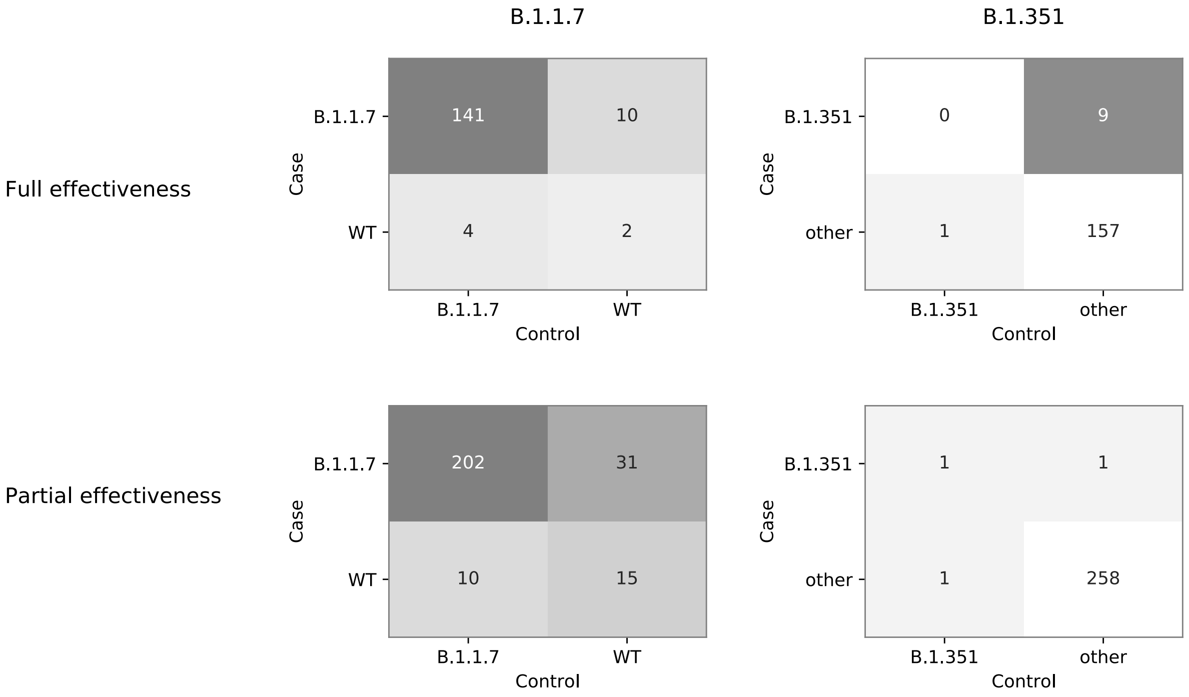


Scenario that most disrupts our results:

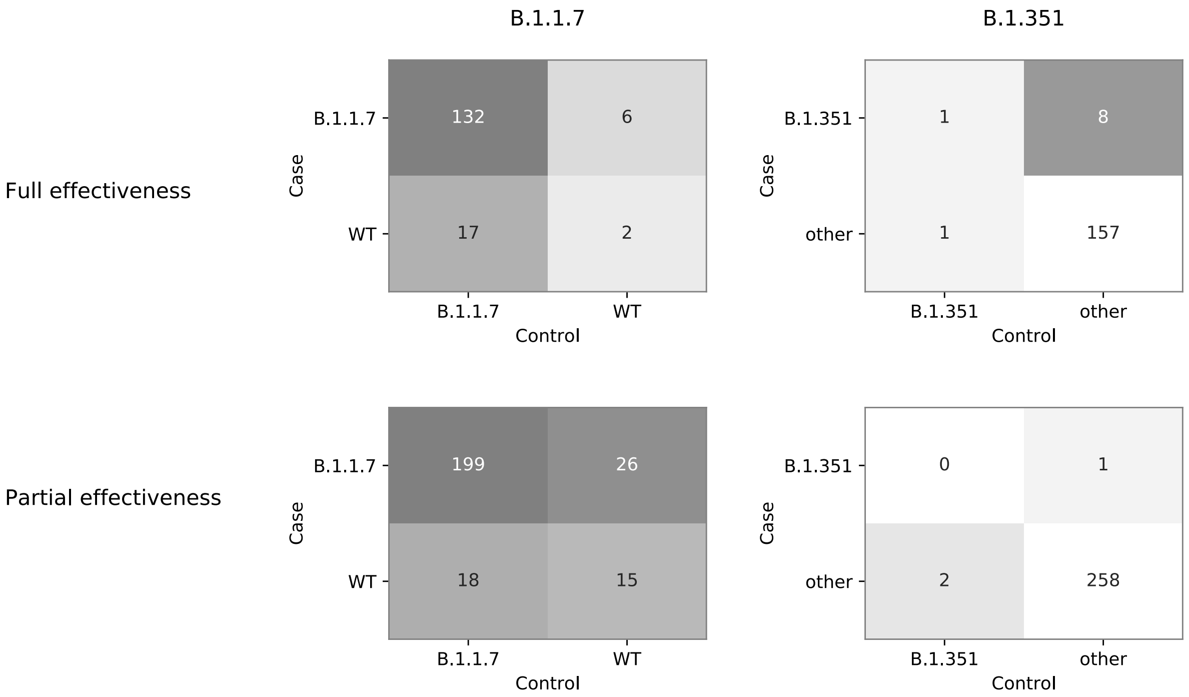


**Figure S3. Sequences whose match (case or control) were not sequenced**. (A) Counts of sequences based on their category (case/control and FE/PE) and variant assignment. (B) We theoretically assign the sequences into the two most extreme scenarios: a scenario that most strengthens our original results as reported in the main text (upper panel), versus a scenario that is most disruptive to the original results (lower panel). Under both scenarios, our results on B.1.351 remain unchanged, with the p-value of the Mcenmar test dropping to 0.01. Results on B.1.1.7 remain qualitatively similar. Under the strengthening scenario the McNemar p-value drops to $7x10^{-4}$, but under the disruptive scenario the McNemar p-value rises to 0.15. We speculate that the true scenario is most likely somewhere in between both extremes.

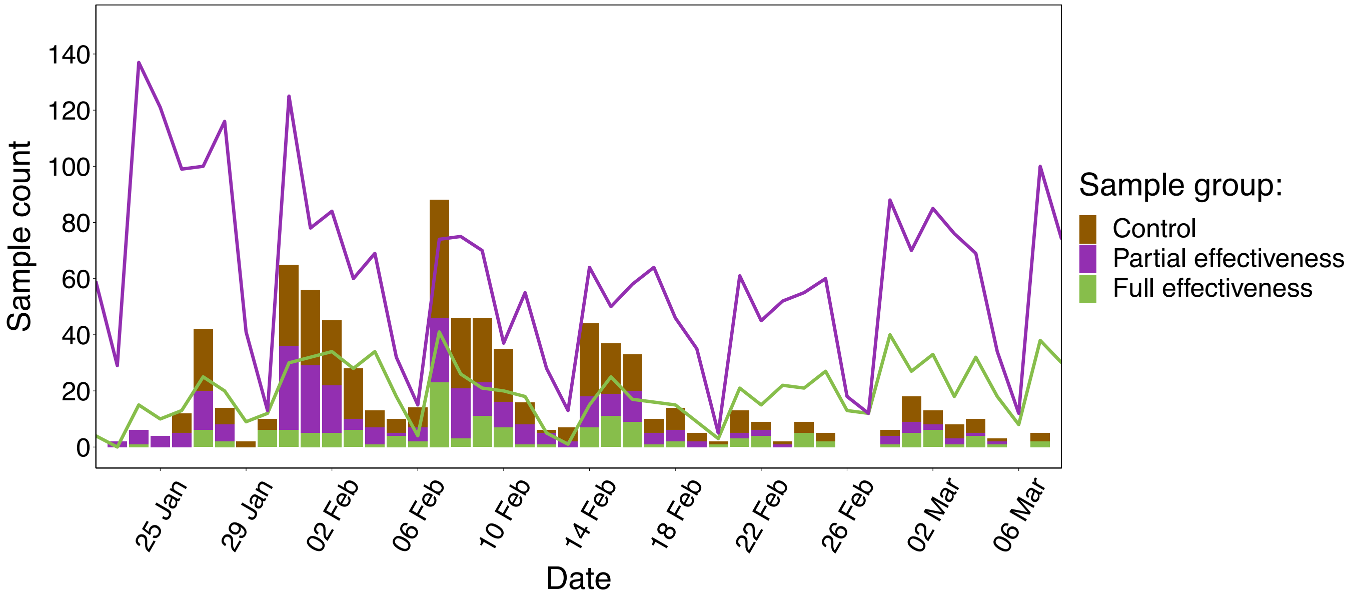


**Fig. S4. Numbers of daily vaccinated carriers.** Data are separated by effectiveness (full effectiveness and partial effectiveness, as defined in the main text) from the six major CHS testing labs located throughout Israel. The bars represent the daily numbers of vaccinees and controls that were successfully sequenced.
